# Free fatty acids accelerate β-cell death in type 1 diabetes

**DOI:** 10.1101/2024.09.16.24313433

**Authors:** Emily C. Elliott, Soumyadeep Sakar, Lisa Bramer, Meagan Burnet, Young-Mo Kim, Xiaoyan Yi, Igor L. Estevao, Marian Rewers, Xiaolu A. Cambronne, Kendra Vehik, Rafael Arrojo e Drigo, Thomas O. Metz, Decio L. Eizirik, Bobbie-Jo M. Webb-Robertson, Raghavendra G. Mirmira, Ernesto S. Nakayasu

**Author notes:** Correspondence (to E.S.N.). Equally contributing authors (Emily C. Elliott, Soumyadeep Sakar).

## Abstract

Type 1 diabetes (T1D) results from autoimmune destruction of the insulin producing pancreatic β cells. The body’s lipid metabolism is strongly regulated during this process but there is a need to understand how this regulation contributes to the β-cell death. Here, we show that fatty acids are released from plasma lipoproteins in children during islet autoimmunity, prior to T1D onset. These fatty acids (FFAs) enhanced cytokine-mediated apoptosis in cultured insulin-producing cells by downregulating the production of nicotinamide adenosine dinucleotide (NAD) via its salvage pathway, as well as deregulated central carbon metabolism and impaired levels of ATP. Downregulation of the NAD salvage pathway and central carbon metabolism enzymes were further observed during T1D development, supporting that the pathways for NAD and energy production are compromised *in vivo*. Our findings show that fatty acids are released during islet autoimmunity, accelerating disease development through impaired NAD metabolism.

## Introduction

Type 1 diabetes (T1D) affects an estimated 8.4 million people worldwide and reduces the life expectancy of those affected by over a decade^1-3^. The disease results from the progressive loss of insulin-producing β cells of the pancreas via an autoimmune response, leading to the inability to control blood glucose levels^4,5^. Selected human leukocyte antigen (*HLA*) genotypes are risk factors, but only 5% of individuals carrying those genotypes eventually develop T1D^6-8^, suggesting the contribution of additional biotic and environmental factors^9^. The body metabolism may be one of these factors as many of its aspects have been associated with T1D development^9-12^. However, little is known about how specific metabolic pathways mechanistically contribute to disease development.

Among the metabolic processes, the body lipid metabolism is one of the most regulated during T1D development^10,12-14^. Plasma lipoproteins are regulated in children even prior to the onset of islet autoimmunity (herein referred to as seroconversion and clinically diagnosed by the appearance of circulating islet autoantibodies)^15^. There is also a decrease in the levels of plasma phosphatidylcholine and triacylglycerols during T1D development^10^. Similarly, in human islets treated with the pro-inflammatory cytokines IL-1β and IFN-γ that mimic the insulitis environment, there is a decrease in triacylglycerol levels^16^. Other lipids, such as short-chain fatty acids, drive inflammation of the intestine, increases intestinal permeability, and augments the exposure to putative dietary antigens^17^. Conversely, diets enriched in omega-3 fatty acids improve intestinal integrity and reduce the onset of T1D in mice^18,19^. Omega-3-rich diet can also diminish the risk of children to develop islet autoimmunity^20^. Mechanistically, omega-3 fatty acids reduce apoptosis of insulin-producing cells by downregulating the methylation of the *ADPRHL2* gene and releasing the expression of its product, the anti-apoptotic factor ARH3.

An open question is how these regulations in the lipid metabolism contribute to T1D development. Here, we hypothesize that downregulations of triacylglycerols (in both plasma and islets) and phosphatidylcholines (in plasma) have a role in β-cell death. To test this hypothesis, we first analyzed lipidomics and proteomics data from human plasma and islets to determine possible regulatory mechanisms of these lipid classes. We also performed experiments in MIN6 insulin-producing cells and analyzed single-cell transcriptomic data to study the downstream consequences of triacylglycerol and phosphatidylcholine downregulation in T1D development. Our data show a mechanism by which lipids accelerate T1D development.

## Material and Methods

### Data from a longitudinal cohort of individuals at high risk of developing T1D

*TEDDY Study and lipidomics dataset*. Six clinical research centers - three in the USA (Colorado, Georgia/Florida, Washington State), and three in Europe (Finland, Germany, and Sweden) participated in a population-based HLA screening of newborns between September 1, 2004 and February 28, 2010^21,22^. The HLA high-risk genotypes identified from the general population (89%) and for infants with a first-degree relative with T1D were previously described^23^. Children enrolled (n=8,676) were prospectively followed from three months of age to 15 years with study visits that include a blood draw every three months until four years and every three or six months thereafter depending on autoantibody positivity. A nested-matched case-control study was conducted through risk-set sampling using metadata and autoantibody sample results as of May 31, 2012, as previously detailed^24^. Additional matching by the clinical center, sex, and family history of T1D (general population or first-degree relative) were included as criteria in the selection of controls. The sample selection was determined by the Data Coordinating Center (University of South Florida Health Informatics Institute, Tampa, FL) without the lab knowing the case-control status. The characteristics of the donors are listed in **Supplemental Table 1**. Plasma lipidomics data were reanalyzed from the datasets described in previous publications^25-27^. The effect of sample analysis order on quantified lipidomics data was removed by applying Systematic Error Removal using Random Forest (SERRF)^28^. All missing values were replaced with “NA”, and data were log2 transformed. For each time point and lipid combination, a paired *t*-test, accounting for case-control pairs, was conducted, and a Benjamini-Hochberg false-discovery rate correction was applied to the resulting p-values^29^.

### GC-MS analysis

Human islets treated with 50 U/mL human IL-1β + 1000 U/mL human IFN-γ (cytokine cocktail 1 or “CT1”) for 24 h were collected as previously described^30^. The Pacific Northwest National Laboratory institutional review board deemed the project to not be human subject research and waived full ethical review. The characteristics of the donors are listed in **Supplemental Table 2**. The data from this experiment were subsequently integrated with proteomics and lipidomics data published elsewhere^16,30^. Samples from either these human islets or MIN6 cells treated as described above were submitted to simultaneous metabolite, protein, and lipid extraction (MPLEx)^31^. Metabolites were treated with 30 mg/mL methoxyamine in pyridine for 90 min at 37 °C with shaking, and derivatized with N-methyl-N-(trimethylsilyl)trifluoroacetamide (MSTFA) (Sigma-Aldrich) with 1% trimethylchlorosilane (TMCS) (Sigma-Aldrich) at 37 °C with shaking for 30 min^32^. Derivatized metabolites were analyzed using an Agilent GC 7890A equipped with an HP-5 MS column (30 m × 0.25 mm × 0.25 μm; Agilent Technologies, Santa Clara, CA) and coupled with a single quadrupole MSD 5975C (Agilent Technologies). Samples were injected into a splitless port set at 250°C with an initial oven temperature of 60 °C. After 1 min the temperature was increased to 325°C at a rate of 10°C/min, and finished with a 5-min hold at 325 °C. The data files were calibrated with external calibration of fatty acid methyl ester and converted to retention time indexes relative to the elution of the fatty acid methyl esters. Molecules were identified by matching experimental retention indices and/or mass spectra to an in-house augmented version of FiehnLib, NIST20, Wiley 11^th^ edition, and MS-DIAL databases (https://systemsomicslab.github.io/compms/index.html)^33,34^. Significant differences in metabolite relative abundances were determined by Student’s *t*-test. The central carbon metabolism pathway was plotted along with each metabolite relative abundance using Vanted^35^.

### Cell culture

MIN6 cells were maintained at 37 ºC in a 5% CO_2_ atmosphere in DMEM containing 4.5 g/L each of D-glucose and L-glutamine, 10% fetal bovine serum, 100 units/mL penicillin, 100 μg/mL streptomycin, and 50 μM 2-mercaptoethanol. For cell assay experiments, cells were treated at 80% confluency with a cytokine cocktail (CT2: 100 ng/mL mouse IFN-γ: R&D, cat#485-MI-100, 10 ng/mL mouse TNF-α: R&D, Cat#410-MT-010, and 5 ng/mL mouse IL-1β: R&D, cat #401-ML-005) for 24 h. Cells were treated concurrently with various concentrations of palmitate (Cayman, CAS#10006627), 10 µM NAMPT inhibitor FK866 (MCE, Cat. No.: HY-50876), 200 µM hexokinase inhibitor lonidamine (Tocris, Cat. No. 1646), or 100 µM nicotinamide riboside (NR) (MCE, HY-123033A). The concentration of NAMPT inhibitor, hexokinase inhibitor, and NR was determined by a dose-response curve, and the concentration with maximum effect was chosen. Palmitate stock was prepared in ethanol and diluted in 0.5% delipidated bovine serum albumin to keep it in solution.

### Proteomics analysis

MIN6 cells were lysed in 50 mM Tris-HCl containing 8 M urea and 10 mM dithiothreitol and incubated for 1 h at 37 ºC with shaking at 800 rpm. Iodoacetamide was added to a final concentration of 40 mM from a 400-mM stock, and the sample was incubated for 1 h in the dark at 21 ºC. The reaction mixture was diluted 8-fold with 50 mM Tris-HCl, and 1 M CaCl_2_ was added to a final concentration of 1 mM. Proteins were digested overnight at room temperature using sequencing-grade trypsin (Promega) at an enzyme-to-protein ratio of 1:50. Digested peptides were desalted by solid-extraction in C18 cartridges (Discovery, 50 mg, Sulpelco) following manufacturer recommendations, and dried in a vacuum centrifuge. Peptides were analyzed using a Waters NanoAquity UPLC system coupled with a Q-Exactive mass spectrometer, as previously described^36^. The data were processed with MaxQuant software (v.1.5.5.)^37^ using the mouse reference proteome database from UniProt Knowledge Base (downloaded on 08/31/2023). Protein N-terminal acetylation and oxidation of methionine were set as variable modifications, while cysteine carbamidomethylation was set as a fixed modification. The software’s default mass shift tolerance was used. Statistical analysis and heatmap were done using Perseus^38^. MaxLFQ intensity was used to quantify protein, being log2 transformed, median normalized, and having the missing values were imputed with Gaussian distribution^39^. Student’s *t*-test was used to determine statistically different proteins. Functional-enrichment analysis was done for the statistically significant proteins using DAVID^40^, using all the predicted mouse genes as the background.

### Apoptosis, NAD, and ATP assays

Apoptosis, NAD, and ATP levels were measured with Caspase-Glo, NAD/NADH-Glo, and CellTiter-Glo® 2.0, respectively, following the manufacturer-provided protocols (Promega Cat# G8092, G9071, and G9242). MIN6 cells were treated with palmitate, FK866, lonidamine, or NR, and CT2 for 24 h. The appropriate amount of the assay reagent was added to the wells. The contents were gently mixed for 30 s and luminescence was read for 3 h every 30-min interval using a plate reader (Synergy HT, BioTek). The time point with the highest signal was selected for analysis. All statistical analysis and data visualization were performed using GraphPad Prism 9 (Version 9.4.1 (458)).

### Single cell RNA seq analysis

The raw single-cell RNA-seq data files (FASTQ, 10X Genomics) of human islets^41^ were downloaded from the Human Pancreas Analysis Program (HPAP) data portal (https://hpap.pmacs.upenn.edu/) and processed with Cell Ranger (v6.1.2). The clinical characteristics of the donors are listed in **Supplementary Table 3**. Data were processed following 10X Genomics recommendations for quality control, read alignment to the reference genome (hg38), barcode processing, and molecule counting. Initial clustering was performed using Seurat software (v4.1.1), followed by decontamination of background mRNA using SoupX (v1.6.1); key genes (*INS, GCG, SST, TTR, IAPP, PYY, KRT19, TPH1*) representing the major cell types were identified in the initial clustering. The adjusted gene expression counts were further processed in Seurat for additional filtering. Potential doublets were identified and removed using scDblFinder (v3.16). Cells were filtered based on the following criteria: nFeature_RNA > 1,000 and < 9,000, mitochondrial gene percentage < 15%, and nCount < 100,000. Following the steps outlined above, we obtained 91,047 single cells for downstream analysis. Normalization of counts was done using the R package scTransform, which adjusts for library size variations per cell. Mitochondrial gene variation was also regressed out, as it indicates cell state. We selected the top 3,000 variable genes to perform principal component analysis (PCA). Data integration was conducted using Harmony (v0.1.1) with the top 50 PCA components, accounting for each sample and reagent kit batches as confounding factors. The integrated components were used to construct a Uniform Manifold Approximation and Projection (UMAP) embedding of the 91,047 cells. Finally, cell types were annotated with scSorter (v0.0.2) based on known marker genes. ExcelChIP-Seq data visualization was performed using IGV software.

## Results

### Pro-inflammatory cytokines induce the release of palmitate in human islets

We have previously shown that triacylglycerols were consistently downregulated in 3 insulitis models: islets from NOD mice in the prediabetic stage, and EndoC-βH1 cells and human islets treated with a cocktail of pro-inflammatory cytokines (CT1: 50 U/mL IL-1β and 1000 U/mL IFN-γ)^16^. To further study this phenotype, we analyzed our previously published proteomics dataset from the same human islets^30^ and found that endothelial lipase LIPG was increased 1.4-fold with CT1 treatment (**Fig. 1A**), suggesting a possible mechanism. A gas chromatography-mass spectrometry analysis of the same human islets treated with CT1 showed that palmitate was increased 1.7-fold with CT1 treatment (**Fig. 1B**). These data show that CT1 induces the digestion of TGs into free fatty acids (FFAs).

**Figure 1:**
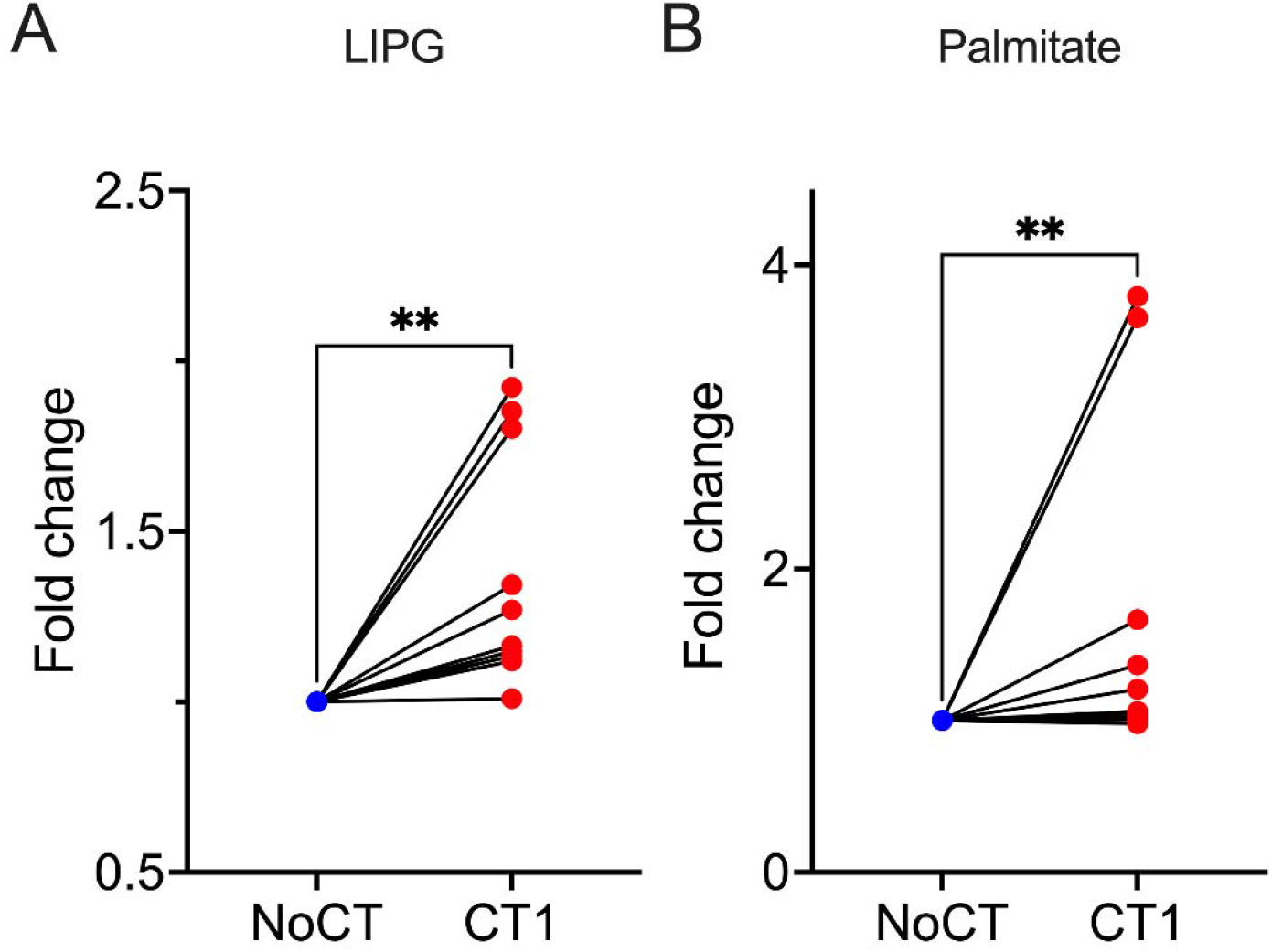
Profiles of endothelial lipase LIPG (A) and free palmitate (B) in human islets treated with cytokines IL-1β and IFN-γ (CT1) for 24 h. LIPG was measured by proteomics analysis while palmitate by gas chromatography-mass spectrometry analysis. NoCT – untreated control. ** Paired Student’s *t*-test p ≤ 0.01.

### Increase of free fatty acid levels in plasma from donors with islet autoimmunity

We investigated if TG digestion and fatty acid release also occurs *in vivo*, during T1D development. As LIPG is a major plasma lipoprotein lipase, we search these signatures on the plasma lipidomics data of the nested case-control 1 study (NCC1) from The Environmental Determinants of Diabetes in the Young (TEDDY) study (**Supplemental Table 4**). In TEDDY NCC1, samples were collected every 3 months from individuals carrying the high-risk HLA alleles from birth until the age of 6 years. We included data from all 94 children who progressed to clinical T1D and from their matched controls. The lipidomics data showed a decrease in the levels of 18 of the 127 phosphatidylcholine species (14%) and 14 of the 100 triacylglycerol species (14%) 3 months post-seroconversion (**Fig. 2A**). The phosphatidylcholine and triacylglycerol intermediate cleavage products lysophosphatidylcholines and diacylglycerols, respectively, displayed a downregulation trend with 3 out of their 32 species (10%) significantly reduced (**Fig. 2B**). Among the FFAs, 2 and 4 of their 31 species (19%) were upregulated at the time of and 3 months post-seroconversion, respectively (**Fig. 2C**). The downstream products from FFAs, acyl-carnitines, had 3 and 7 of their 17 species (59%) upregulated at the time of and 3 months post-seroconversion, respectively (**Fig. 2C**). This suggests that the extent of fatty acid release is higher than the one observed based on FFA abundance alone.

**Figure 2.**
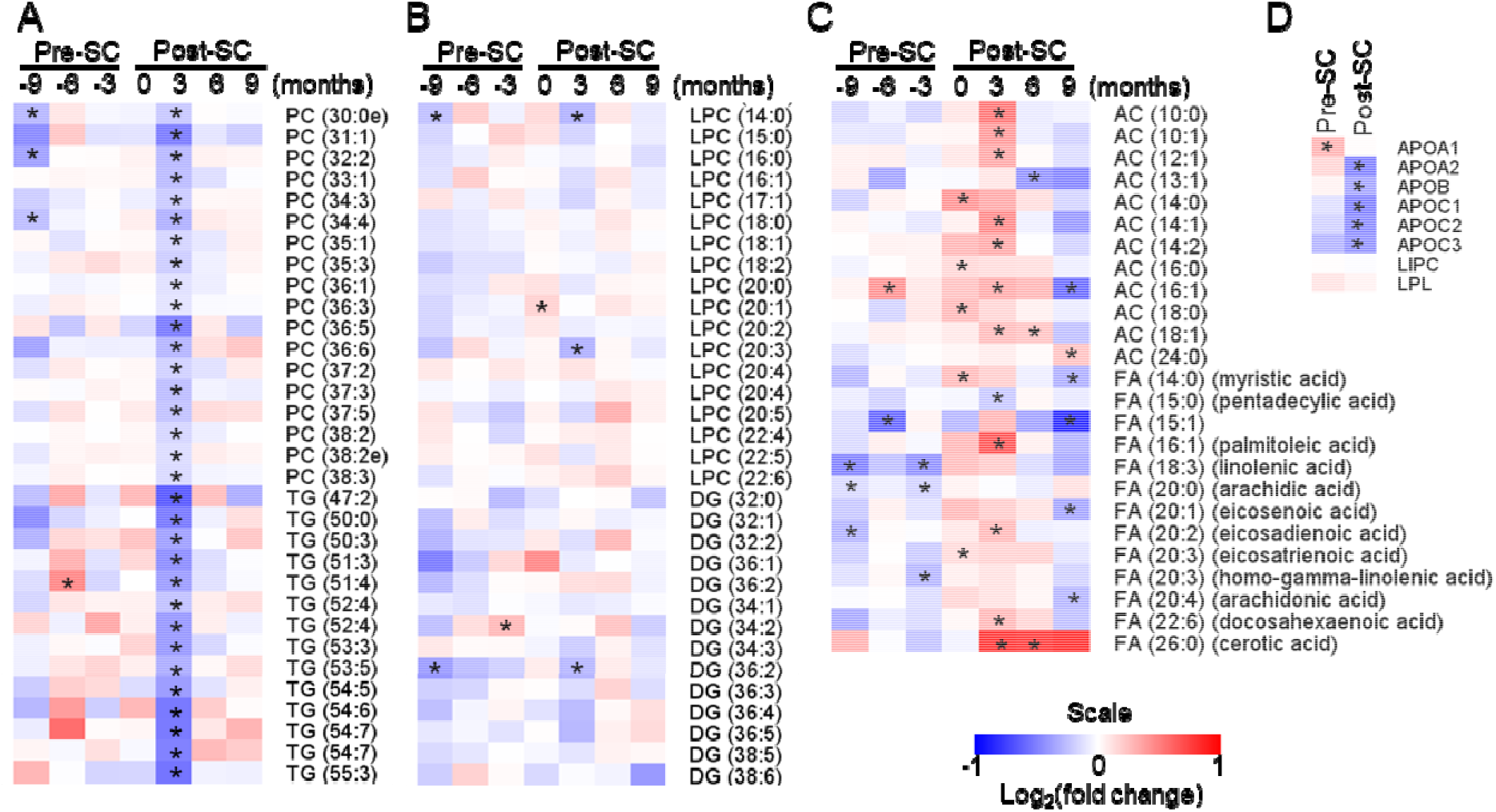
Plasma lipid and lipoprotein profile on during T1D development. Lipid and protein profiles were obtained from lipidomics and proteomics measurements of the TEDDY nested case-control study 1 (NCC1), which comprises of time point sample collections from children up to the age of 6 years. (A) Heatmap of representative, significant species of phosphatidylcholines (PC) and triacylglycerols (TG). (B) Heatmap of all quantified intermediates of PC and TG, lysophosphatidylcholine (LPC) and diacylglycerol (DG), respectively. (C) Heatmap of representative, significant species of PC and TG digestion products, acyl-carnitines (AC) and (free) fatty acids (FA), respectively. Additional abbreviations: ApoA1 – apolipoprotein A1, ApoA2 – apolipoprotein A2, ApoC1 – apolipoprotein C1, ApoC2 – apolipoprotein C2, ApoC3 – apolipoprotein C3, LIPC – hepatic lipase, LPL – lipoprotein lipase, Pre-SC – pre-seroconversion, Post-SC – post-seroconversion. *Student’s *t*-test ≤ 0.05.

We also examined the abundance of key lipoprotein subunits and lipases in TEDDY NCC1 proteomics data^15^. Apolipoprotein ApoA1 was upregulated pre-seroconversion, whereas ApoA2, ApoB, ApoC1, ApoC2 and ApoC3 were downregulated post-seroconversion (**Fig. 2D**). Regarding lipases, the lipoprotein lipase LPL and the hepatic lipase LIPC had similar abundances pre- and post-seroconversion compared to the control group (**Fig. 2D**). LIPG was not detected in the analysis.

Overall, these data suggest a digestion of phosphatidylcholines and triacylglycerols, leading to the release of FFAs. These changes are accompanied by reduction in multiple lipoprotein subunits.

### Palmitate enhances pro-inflammatory cytokine-mediated apoptosis

To study if the FFAs have a role in β-cell death, we measured apoptosis in MIN6 cells treated with a cytokine cocktail (CT2: 100 ng/mL IFN-γ, 10 ng/mL TNF-α, and 5 ng/mL IL-1β) for 24 h in combination with a variety of fatty acids at 400 μM: oleate (18:1), stearate (18:0), palmitate (16:0), arachidonate (20:4), linoleate (18:2), docosahexaenoate (22:6) and eicosapentaenoate (20:5). While stearate, palmitate and linoleate had minimal to no effect on MIN6 cells without cytokines, they synergistically enhanced the cytokine-mediated apoptosis (**Fig. 3A**). Conversely, arachidonate and eicosapentaenoate reduced apoptosis in both cells treated or not treated with CT2 (**Fig. 3A**). As palmitate had the strongest effect and was also released by LIPG in islets, it was chosen for the remaining experiments. We treated MIN6 cells with various concentrations of palmitate (50-800 μM) in combination with CT2 for 24 h and measured apoptosis. The palmitate treatment itself had no effect on β-cell apoptosis up to 400 μM, but concentrations as low as 200 μM significantly enhanced the apoptosis induced by CT2 (**Fig. 3B**). These results showed that FFAs increase cytokine-mediated apoptosis.

**Figure 3.**
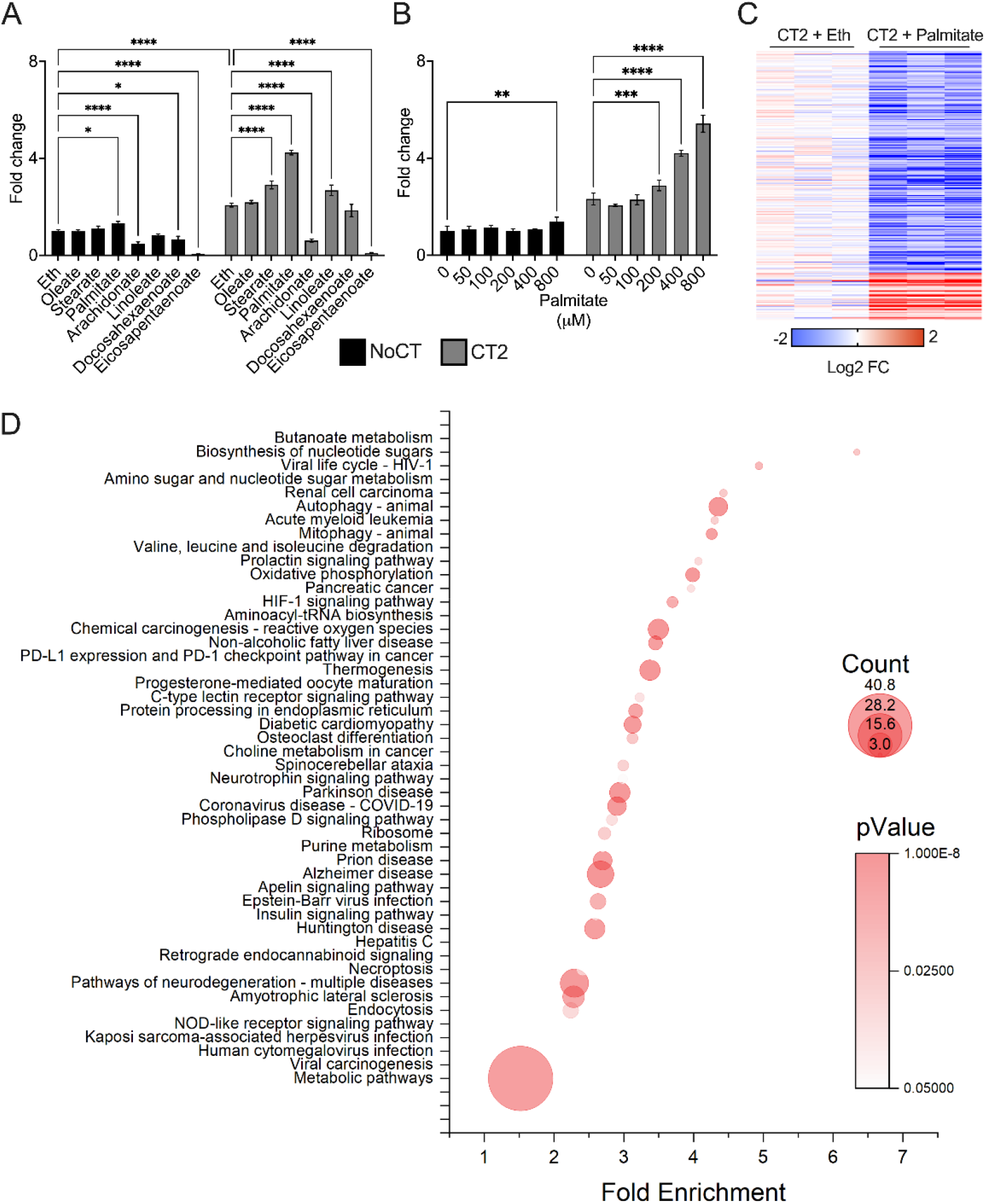
Enhancement of cytokine-mediated apoptosis by palmitic acid. (**A**) MIN6 cells were treated for 24 h with a cytokine cocktail (CT2: 100 ng/mL IFN-γ, 10 ng/mL TNF-α, and 5 ng/mL IL-1β) in combination with various types of fatty acids at a concentration of 400 μM: oleate, stearate, palmitate, arachidonate, linoleate, docosahexaenoate, eicosapentaenoate and apoptosis levels measured by caspase 3/7 activity using a luminescent assay. (**B**) The cells were treated with CT2 in combination with various concentrations of palmitate and apoptosis was measured. (**C**) Heatmap of significant proteins comparing CT2 vs. CT2 + 400 μM palmitate. (**D**) Function enrichment analysis of significant proteins comparing CT2 vs. CT2 + palmitate. Additional abbreviations: NoCT – no cytokine treatment control, Eth – ethanol vehicle control. 2-way ANOVA, Šídák’s multiple comparisons test: *p ≤ 0.05, ** p ≤ 0.01, *** p ≤ 0.001, **** p ≤ 0.0001.

### NAMPT as a regulator of FFA-enhanced cytokine-mediated apoptosis

To identify possible mechanisms of FFA enhancement of cytokine-mediated apoptosis, we performed a proteomics analysis of MIN6 cells treated with CT2 + 400 μM palmitate, leading to the identification of 6049 proteins (**Supplemental Table 5**). The CT2 treatment itself regulated the levels of 631 proteins (86 upregulated and 545 downregulated), while the palmitate treatment itself regulated the levels of 262 proteins (10 upregulated and 252 downregulated) (**Supplemental Table 6-7**). The CT2 + palmitate vs. CT2 comparison resulted in abundance changes of 261 proteins (45 and 216 proteins upregulated and downregulated, respectively) (**Fig. 3C, Supplemental Table 8**). A functional-enrichment analysis of this comparison identified 48 pathways to be significantly enriched among the regulated proteins (**Fig. 3D**). This included metabolic pathways (e.g. oxidative phosphorylation, amino sugar and nucleotide sugar metabolism, insulin signaling pathway), cell-death pathways (e.g. necroptosis, NOD-like receptor signaling pathway) and immune response pathways (e.g. PD-L1 expression, human cytomegalovirus infection).

We next investigated possible mechanisms of apoptosis enhancement by the CT2 + palmitate treatment. In the proteomics analysis, the NOD-like receptor signaling pathway was the one directly involved in apoptosis among those regulated when comparing CT2 + palmitate vs. CT2 conditions (**Fig. 3D**). One standout member of this pathway was nicotinamide phosphoribosyltransferase, NAMPT, whose levels were upregulated 4.5-fold in cells treated with CT2 and diminished by 0.6-fold following addition of palmitate to the CT2 treatment (**Fig. 4A**). NAMPT is also the rate-limiting enzyme of the intracellular nicotinamide adenosine dinucleotide (NAD) salvage biosynthesis pathway. To test the role of NAMPT depletion in enhancing CT2-mediated apoptosis, we treated MIN6 cells with its inhibitor FK866 in combination with CT2. The inhibition of NAMPT induced a phenotype similar to that of palmitate-induced enhancement of CT2-mediated apoptosis, which was abolished by supplementing the cells with nicotinamide riboside (NR) (**Fig. 4B**), a metabolite downstream from NAMPT in the NAD salvage pathway. We next measured the levels of NAD in FK866-treated cells, which confirmed that FK866 causes depletion of NAD and that its production was rescued by the addition of NR **(Fig. 4C)**. We treated MIN6 cells with CT2 + palmitate in combination with NR and measured both apoptosis and NAD levels. The NR supplementation failed to rescue both the apoptosis enhancement by palmitate (**Fig. 4D**) and the recovery of cellular NAD levels (**Fig. 4E**). These data show that palmitate enhances cytokine-mediated apoptosis via downregulation of NAMPT and depletion of NAD levels. However, NR supplementation fails to rescue the cells from apoptosis, suggesting that additional factors are involved.

**Figure 4.**
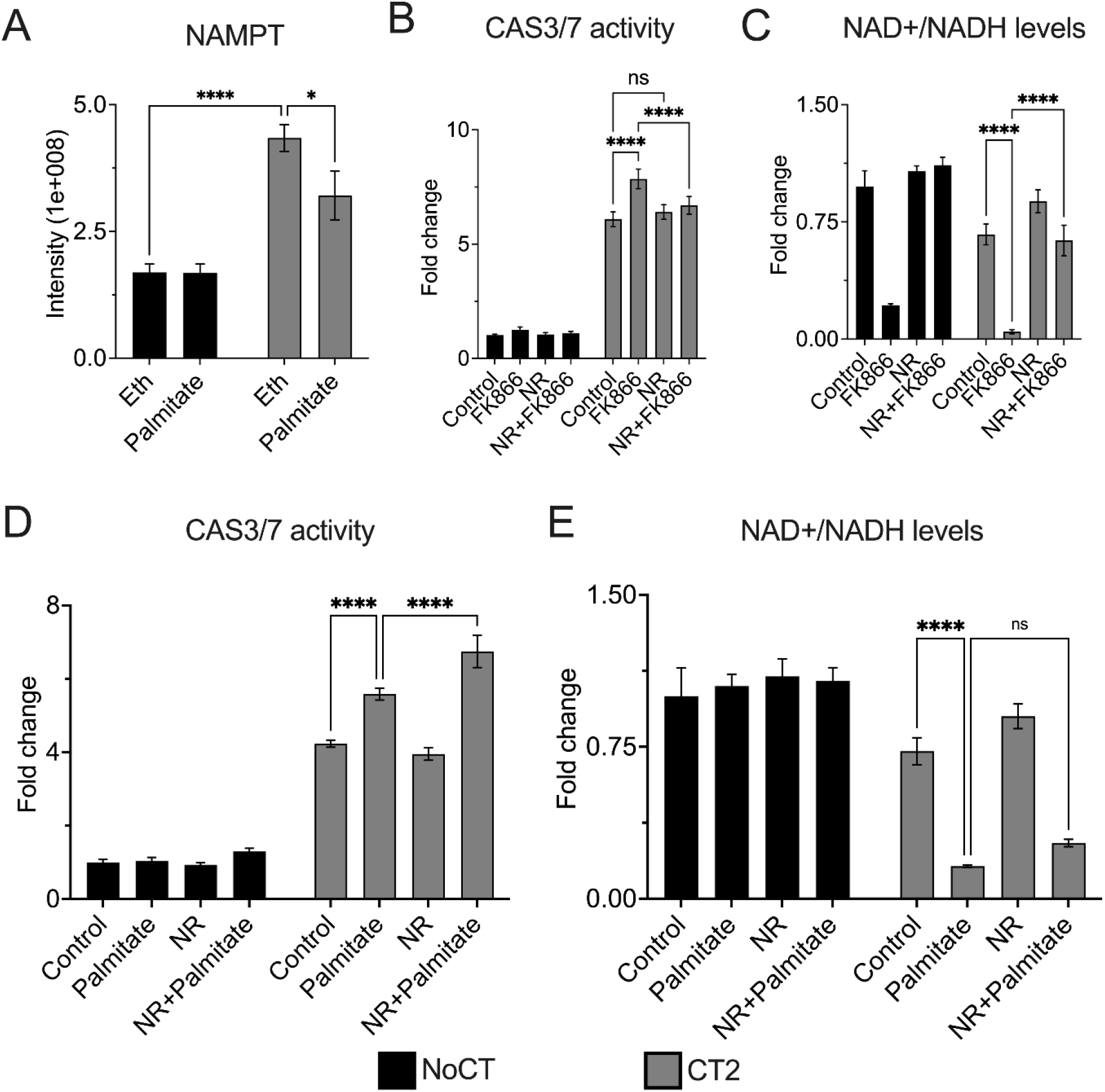
Regulation of NAD metabolism in cytokine-mediated apoptosis by palmitate. MIN6 cells were treated with cytokine cocktail (CT2: 100 ng/mL IFN-γ, 10 ng/mL TNF-α, and 5 ng/mL IL-1β) in combination with 400 µM palmitate. (**A**) Levels of NAMPT measured by proteomics analysis. Levels of apoptosis (measured by caspase 3/7 activity) (**B**) and NAD (**C**) in cells treated with CT2 in combination with 10 µM NAMPT inhibitor FK866 and 100 µM nicotinamide ribose (NR). Levels of apoptosis (measured by caspase 3/7 activity) (**D**) and NAD (**E**) in cells treated with CT2 + 400µM palmitate in combination with 100 µM NR. Additional abbreviations: NoCT – no cytokine treatment control, Eth – ethanol vehicle control. 2-way ANOVA, Šídák’s multiple comparisons test: *p ≤ 0.05, ** p ≤ 0.01, *** p ≤ 0.001, **** p ≤ 0.0001.

### Deregulation of the central carbon metabolism by cytokines + palmitate reduces cellular ATP levels

We next investigated possible additional factors that impair NR from rescuing cells from the cytokine-mediated apoptosis enhanced by palmitate. The insulin receptor signaling pathway, which regulates glycolysis, was one of the pathways identified as downregulated in the proteomics analysis comparing the CT2 + palmitate vs. CT2 conditions (**Fig. 3C**). Eight proteins of this pathway were downregulated, including the glycolysis rate-limiting enzyme hexokinase 1 (HK1) (**Fig. 5A**). In MIN6 cells, the CT2 treatment alone decreased the level of HK1 by 1.6-fold, but the combination of CT2 + palmitate reduced the levels of HK1 by 33% (**Fig. 5B**). To investigate possible consequences of this regulation on central carbon metabolism, we performed a GC-MS metabolomics analysis of MIN6 cells treated with the combination of CT2 + palmitate for 8 h. The palmitate treatment alone only reduced the levels of lactate (**Fig. 5C**), while CT2 treatment alone induced a trend for increasing the glycolytic activity with elevated but not significant pyruvate concentration (**Fig. 5C**). The combined CT2 + palmitate treatment led to an increase in phosphoenolpyruvate and reduction in lactate levels compared to the control (**Fig. 5C**). Downstream, there was an increase in the late TCA cycle metabolites α-ketoglutarate, malate and fumarate comparing the CT2 + palmitate vs. the CT2 alone conditions (**Fig. 5C**). There is an increase in glutamate and glutamine (**Fig. 5C**), suggesting an overflow from the central carbon metabolism towards the amino acid metabolism.

**Figure 5.**
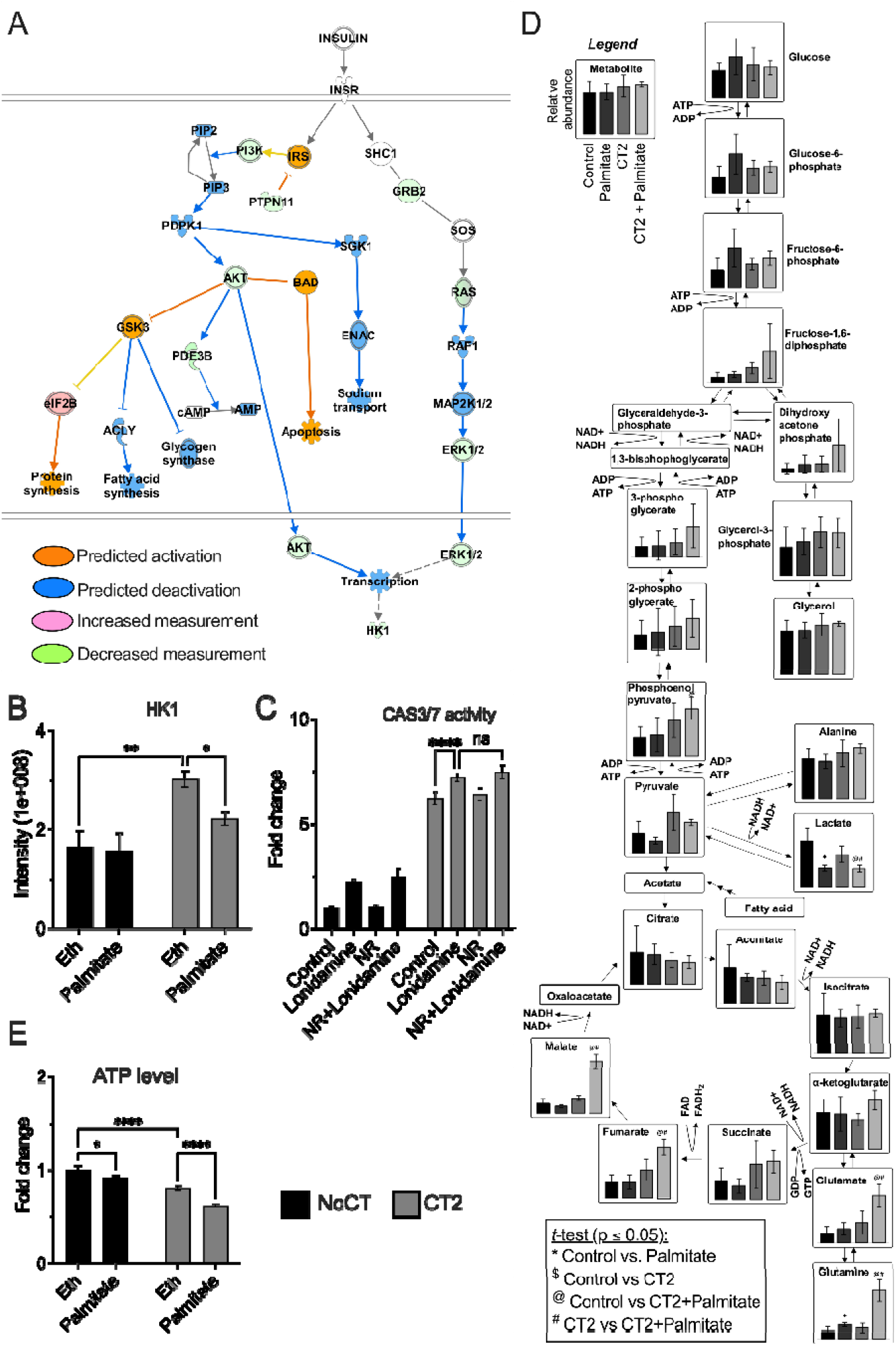
Regulation of the central carbon metabolism by the cytokine cocktail (CT2: 100 ng/mL IFN-γ, 10 ng/mL TNF-α, and 5 ng/mL IL-1β) + palmitate. (**A**) Enrichment of palmitate + CT2-regulated proteins onto the insulin receptor signaling pathway using IPA. (**B**) Hexokinase 1 HK1 protein level measured by proteomic analysis. Levels of apoptosis (measured by caspase 3/7 activity) (**C**) in MIN6 cells treated with CT2 with or without 400 μM palmitate in combination with 200 µM hexokinase inhibitor lonidamine and 100 µM nicotinamide riboside (NR). (**D**) Level of various metabolites from the central carbon metabolism measured using GC-MS in MIN6 cell treated with CT2 + 400 µM palmitate. (**E**) Level of ATP measured in MIN6 cell treated with CT2 + 400 µM palmitate. Additional abbreviations: NoCT – no cytokine treatment control, Eth – ethanol vehicle control. T-test (**D**), 2-way ANOVA, Šídák’s multiple comparisons test (**B, C**, & **E**): ** p ≤ 0.01, *** p ≤ 0.001, **** p ≤ 0.0001.

Next, we measured the levels of ATP to determine how these changes in the central carbon metabolism affect energy production. The ATP level was reduced by 8% in cells treated solely with palmitate and by 20% in cells treated with CT2. The CT2 + palmitate treatment reduced ATP by 38% (**Fig. 5E**), showing that cellular energy production is compromised. To test the role of HK1 in the regulation of the CT2 + palmitate enhanced apoptosis, we treated the cells with the HK1 inhibitor lonidamine. The treatment enhanced the CT2-mediated apoptosis in MIN6 by 1.25-fold (**Fig. 5D**). However, a similar increase was also observed in cells not treated with cytokines (1.4-fold increase) (**Fig. 5D**), suggesting that HK1 is not responsible for the enhancement of cytokine-mediated apoptosis by palmitate. These results show that downregulation of HK1 is not responsible for enhancement of cytokine-mediated apoptosis by palmitate. However, FFA in combination with cytokines affected central carbon metabolism and ATP production, which might contribute to the depletion of NAD as the salvage pathway requires ATP.

### Downregulation of the NAD salvage pathway and central carbon metabolism in β cells during T1D development

To study if the NAD salvage pathway and central carbon metabolism regulation may occur *in vivo* during T1D development, we analyzed the islet single cell RNA seq data from human donors in different stages of T1D development^42^. These data contained quantitative sequencing information from 12,660 β cells, including 4,710 from non-diabetic donors, 4,156 from donors with single islet autoantibodies, 2,521 from donors with multiple islet autoantibodies, and 1,273 from donors with T1D. In the NAD salvage pathway, we found that NAMPT expression is higher in β cells of non-diabetic donors, and significantly reduced in individuals with single or multiple autoantibodies (**Fig. 6A**). In central carbon metabolism, there is a downregulation of 12 of the 20 detected transcripts of the glycolytic pathway in β cells in individuals with one or multiple islet autoantibodies (**Fig. 6B**). After the onset of the disease, the levels of the glycolytic enzymes are slightly higher with the transcripts of 3 enzymes upregulated (**Fig. 6B**). A similar pattern is observed in the TCA cycle, with the expression of 12 enzymes reduced in individuals positive for one or multiple autoantibodies, and 4 enzymes increased after the onset of the disease (**Fig. 6C**). These results show that the expression of the components of the NAD salvage pathway, glycolysis, and TCA cycle are downregulated in β cells during T1D development, supporting our *in vitro* findings.

**Figure 6.**
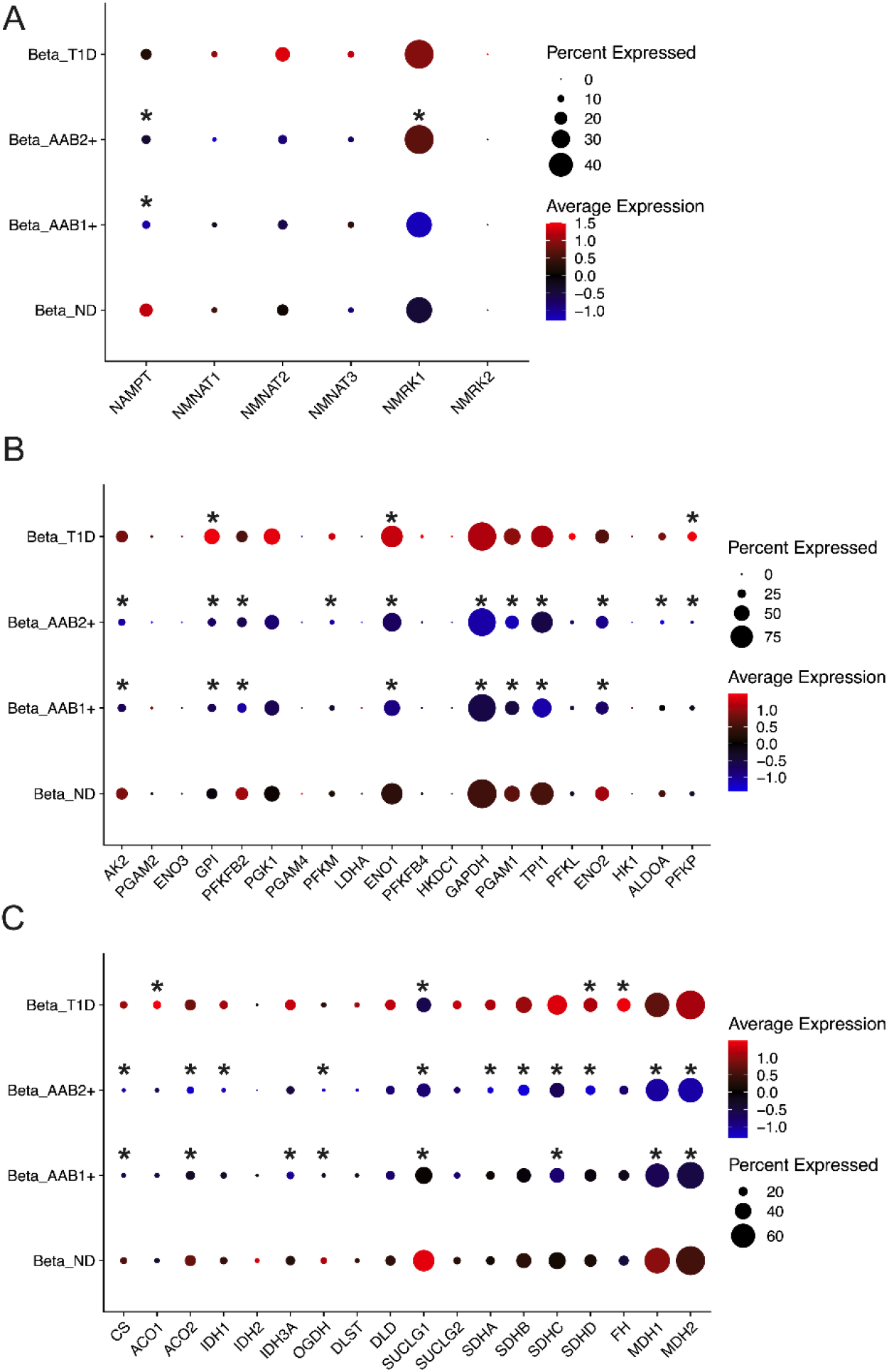
Expression of genes from the NAD salvage pathway, glycolysis/gluconeogenesis and TCA cycle during different stages of type 1 diabetes development. The data were downloaded from the Human Pancreas Analysis Program (HPAP) data portal (https://hpap.pmacs.upenn.edu/) and processed ad described in material and methods. Abbreviations: AAB1+ - individuals seropositive for 1 islet autoantibody, AAB2+ - individuals seropositive for multiple islet autoantibodies, beta – β cells, T1D – type 1 diabetes.

## Discussion

Our current and previous data^16^ show that CT1 reduced the levels of triacylglycerols, which was associated to LIPG upregulation and their digestion into FFAs. In the TEDDY plasma lipidomics data, there is a similar pattern with decrease in triacylglycerols and phosphatidylcholines and an increase in FFAs at 3 months post-seroconversion. Although some of the increases in FFA levels are moderate, we found that concentrations as low as 200 μM of palmitate enhances cytokine-mediated apoptosis. Considering that the basal levels of palmitate in plasma are around 145 μM^44^, even small changes in FFA concentration might have a deleterious effect when combined with the local pro-inflammatory cytokines present during insulitis in a chronic disease.

Regarding specific plasma lipoproteins, ApoA1 and ApoA2 (which were regulated in the proteomics analysis) are major components of HDL and chylomicrons^45^, but LIPG being an HDL lipase^46^, making more likely that high-density lipoprotein (HDL) is involved in the process. HDL is also the particle richest in phospholipids^45^, and is a possible source of the phosphatidylcholines digested after seroconversion. Likewise, HDL decreases after seroconversion but returns to basal levels after T1D diagnosis, suggesting a temporary regulation^47^. ApoB was also reduced after seroconversion in our proteomics analysis. This protein has two major isoforms, the full-length ApoB-100, enriched in low-density lipoprotein (LDL) and very low-density lipoprotein (VLDL), and ApoB-48, enriched in chylomicrons^45^. Unfortunately, our bottom-up proteomics analysis does not distinguish between the two isoforms. Considering that all three particles are rich in triacylglycerols^45^, each could be the source of the triacylglycerols that are digested after seroconversion. During islet autoimmunity, LDL levels remain unaltered but are increased later^47^.

Regarding lipases involved in the fatty acid release, LIPG is a cell surface lipoprotein lipase that digests lipids from HDL and facilitates the uptake of fatty acids by cells^43^. LIPG also helps to dock and internalize HDL, LDL, VLDL particles by cells^43^. LIPC is less likely to be involved as its regulation is mainly via transcription^48^ and its levels in the proteomics analysis remained unaltered. LPL protein levels also remained unaltered, but its activity might be enhanced by the observed reduction in its inhibitors ApoC1 and ApoC3. Low ApoC1 is associated with progression to T1D before the age of 5 years^49^. There is a similar pattern of ApoC1 in the TEDDY NCC1, where its reduction was associate with the development of T1D by the age of 6 years^15^. LIPG and LPL might work sequentially to digest triacylglycerols and phosphatidylcholines, as LIPG mainly cleaves fatty acyl groups at the *sn-1* position into diacylglycerols and lysophosphatidylcholines^43^. These intermediates largely remained unaltered in the process, suggesting that they are being cleaved by the action of additional lipase(s).

Both LIPG and LPL play relevant roles in inflammation. In macrophages, LIPG mediates toll-like receptors 3 and 4 signaling by repressing and inducing the expression of anti- and pro-inflammatory genes, respectively^43^. LPL enhances interferon-γ-induced pro-inflammatory signaling in endothelial cells^50^. Our results show that released FFAs enhance cytokine-mediated apoptosis in MIN6 cells, which involves the depletion of NAD. Pro-inflammatory cytokines induce an upregulation of the NAD *de novo* synthesis pathway in human islets^51^. This pathway promotes an efficient inflammatory response by fueling the cell with NAD^52^, and might provide the NAD needed during insulitis. Our data show that the combination of CT2 + palmitate inhibits the NAD salvage pathway, depleting cellular NAD. In the body, the *de novo* synthesis pathway is particularly active in the liver, while other tissues rely mostly on the salvage pathway^53^. This agrees with the strong impact we observed in cells treated with cytokines + palmitate. In hepatocytes, palmitate downregulates the expression of NAMPT, leading to apoptosis^54^. However, in contrast to our observations, the hepatocyte apoptosis is reversed by the addition of nicotinamide mononucleotide, a metabolite downstream from NAMPT in the NAD salvage pathway^54^. In our experimental condition, the cells are not rescued with NR, another metabolite downstream from NAMPT in the NAD salvage pathway.

We found that the combination of FFAs with cytokines induced major changes cellular central carbon metabolism. The cells treated with cytokines + palmitate had an unproductive accumulation of various glycolytic and TCA cycle intermediates, leading to reduction in cellular ATP levels. This might explain the inability to rescue the cells with NR, as both enzymes of the salvage pathway, NAMPT and NMNAT, require ATP to catalyze their reactions. Therefore, the deregulation in central carbon metabolism further aggravates the impairment in the NAD salvage pathway^55^. Overall, our data show that inflammation leads to a release in FFAs, which aggravates cytokine-mediated apoptosis. This process was associated with the development of T1D at a young age (<6 years). The mechanism involves depletion of cellular NAD levels by impairing its recycling via the salvage pathway, which is further aggravated by deregulation of central carbon metabolism leading to reduced ATP production.

## Competing interests

The authors declare that they have no competing interests.

## Acknowledgments

Part of the work was performed in the Environmental Molecular Sciences Laboratory, a U.S. DOE national scientific user facility at Pacific Northwest National Laboratory (PNNL) in Richland, WA. Battelle operates PNNL for the DOE under contract DE-AC05-76RLO01830. This work was supported by the Catalyst Award from the Human Islet Research Network (HIRN) (to E.S.N) (via U24 DK104162) and by National Institutes of Health, National Institute of Diabetes and Digestive and Kidney Diseases (NIDDK) grants R01 DK138335 (to E.S.N., M.R., B.J.M.W.R and T.O.M.), U01 DK127505 (to E.S.N.), U01 DK127786 (to R.G.M, D.L.E, B.J.M.W.R and T.O.M.), R01 DK060581 (to R.G.M.), R01 DK105588 (to R.G.M.) The TEDDY Study is funded by U01 DK63829, U01 DK63861, U01 DK63821, U01 DK63865, U01 DK63863, U01 DK63836, U01 DK63790, UC4 DK63829, UC4 DK63861, UC4 DK63821, UC4 DK63865, UC4 DK63863, UC4 DK63836, UC4 DK95300, UC4 DK100238, UC4 DK106955, UC4 DK112243, UC4 DK117483, U01 DK124166, U01 DK128847, and Contract No. HHSN267200700014C from the NIDDK, National Institute of Allergy and Infectious Diseases (NIAID), Eunice Kennedy Shriver National Institute of Child Health and Human Development (NICHD), National Institute of Environmental Health Sciences (NIEHS), Centers for Disease Control and Prevention (CDC), and JDRF. This work is supported in part by the NIH/NCATS Clinical and Translational Science Awards to the University of Florida (UL1 TR000064) and the University of Colorado (UL1 TR002535). The content is solely the responsibility of the authors and does not necessarily represent the official views of the National Institutes of Health.

## Data availability

The TEDDY lipidomics data are available at the Metabolomics Workbench repository under project ID PR000950 and doi 10.21228/M8WM4P. The GC-MS data were uploaded into the Open Science Framework under accession number 86u3m (https://osf.io/86u3m/). The proteomics data were deposited into the MassIVE repository, a member of the ProteomeXchange, under accession number MSV000095733.

## Notes

### Competing Interest Statement

The authors have declared no competing interest.

### Author Declarations

The Pacific Northwest National Laboratory institutional review board deemed the project to not be human subject research and waived full ethical review.

